# ARCADE: Enhancing Automated Document Analysis Through Adversarial Multi-Agent Validation

**DOI:** 10.64898/2025.12.21.25342744

**Authors:** Caleb J. Kumar, Hugh Pearson, Che L. Reddy

## Abstract

We present ARCADE (Adversarial Critique Architecture for Document Evaluation), a multi-agent architecture addressing three limitations of traditional retrieval-augmented generation for automated document analysis: incomplete information extraction, shallow analytical depth, and framework paraphrasing. We compared ARCADE against Single-Pass RAG using 95 policy documents (50 National Cancer Control Plans and 45 Cardiovascular Disease plans) evaluated on 36 metrics across six capabilities: Natural Language Understanding, Logical Reasoning, Fluency, Coherence, Factual Grounding, and Synthesis. Results demonstrate three primary improvements. First, ARCADE achieves 100% extraction success compared to 60% for Single-Pass RAG, eliminating systematic failures that affect nearly half of framework dimensions. Second, analytical depth increases substantially (93-150% longer responses) while readability simultaneously improves by 12 points on the Flesch Reading Ease scale, moving from “very difficult” to “difficult” reading level. Third, multiple independent metrics confirm genuine critical evaluation rather than redundancy: 60% score disagreement between methods, 17-30% factual content overlap, and decreased framework semantic similarity. These gains require minimal trade-off: a 9% reduction in lexical diversity suggests more consistent, if slightly repetitive, terminology. These findings establish adversarial multi-agent validation as an effective paradigm for automated document assessment, with clear applications in policy analysis, regulatory compliance, and clinical protocol evaluation.

## 1. Introduction

Automated evaluation of complex documents against structured frameworks is essential across all major sectors and industries. From public policy and healthcare to legal enterprise, organizations must verify that contracts, strategic plans, and protocols align with mandatory standards. Currently, the development, review, and analysis of complex documents is time-consuming, error-prone, and costly—systematic reviews alone average over 1,000 hours.^[1]^

Complex documents are long and structurally heterogeneous, and often multilingual. They contain dense tables, figures, and appendices, making manual review slow and variable. Examples include national health strategies spanning hundreds of pages or complex legal agreements and regulatory submissions.

Current automation relies on retrieval-augmented generation (RAG) with document preprocessing and optical character recognition (OCR) to ground model generations in framework definitions and retrieved evidence.^[2,3]^

However, these approaches show three systematic failures.^[4]^ First, they often paraphrase framework definitions rather than providing original, document-grounded judgments. Second, retrieval-centered pipelines are prone to hallucination, selective citation, and shallow synthesis.^[5,6,20]^ Third, standard RAG is single pass, lacking mechanisms to interrogate reasoning or revise initial outputs.

Improving automated assessment requires new techniques to address these failures. A promising approach lies in critic-style mechanisms, which demonstrate improved reasoning reliability through structured critique and iterative refinement.^[7]^ However, there are no described approaches in the literature to harness critical-style techniques for complex document analysis.

We introduce ARCADE (Adversarial Critique Architecture for Document Evaluation), a three-stage semantic pipeline for automated complex document analysis. ARCADE preserves retrieval grounding but adds adversarial validation: a dedicated critic agent systematically challenges the analytical agent’s scores and reasoning through structured debate cycles.

### 1.1 Contributions

This work makes four primary contributions. First, we introduce an adversarial multi-agent architecture using a dedicated critic agent to improve analysis through structured critique. The architecture implements specific prompting strategies such as devil’s advocate reasoning, evidence gap analysis, semantic coherence assessment, and comparative robustness testing.

Second, we provide a replicable evaluation framework benchmarking Single-Pass RAG against ARCADE using 36 cognitive metrics organized across six capabilities: Natural Language Understanding & Comprehension, Logical Reasoning & Natural Language Inference, Fluency & Grammaticality, Coherence & Discourse Structure, Factual Grounding & Faithfulness, and Synthesis & Creativity. This multidimensional framework systematically assesses the shift from framework paraphrasing to original content generation.

Third, we demonstrate ARCADE’s effectiveness in the analysis of 95 policy documents—50 National Cancer Control Plans (NCCPs) and 45 National Cardiovascular Disease Control Plans (NCVDCPs) plans. These documents exemplify complex document analysis: they are 20-100 pages, structurally heterogeneous, multilingual, dense with tables and figures, and require assessment against a detailed domain-specific framework. Results show that adversarial validation increases analytical depth by 93-150% while improving readability by 12 points.

Fourth, we outline a development roadmap including multi-agent roundtables for structured deliberation, and domain-specific fine-tuning to build critic agent expertise in a specific subject matter.

## 2. Architecture Comparison

**Figure 1.**
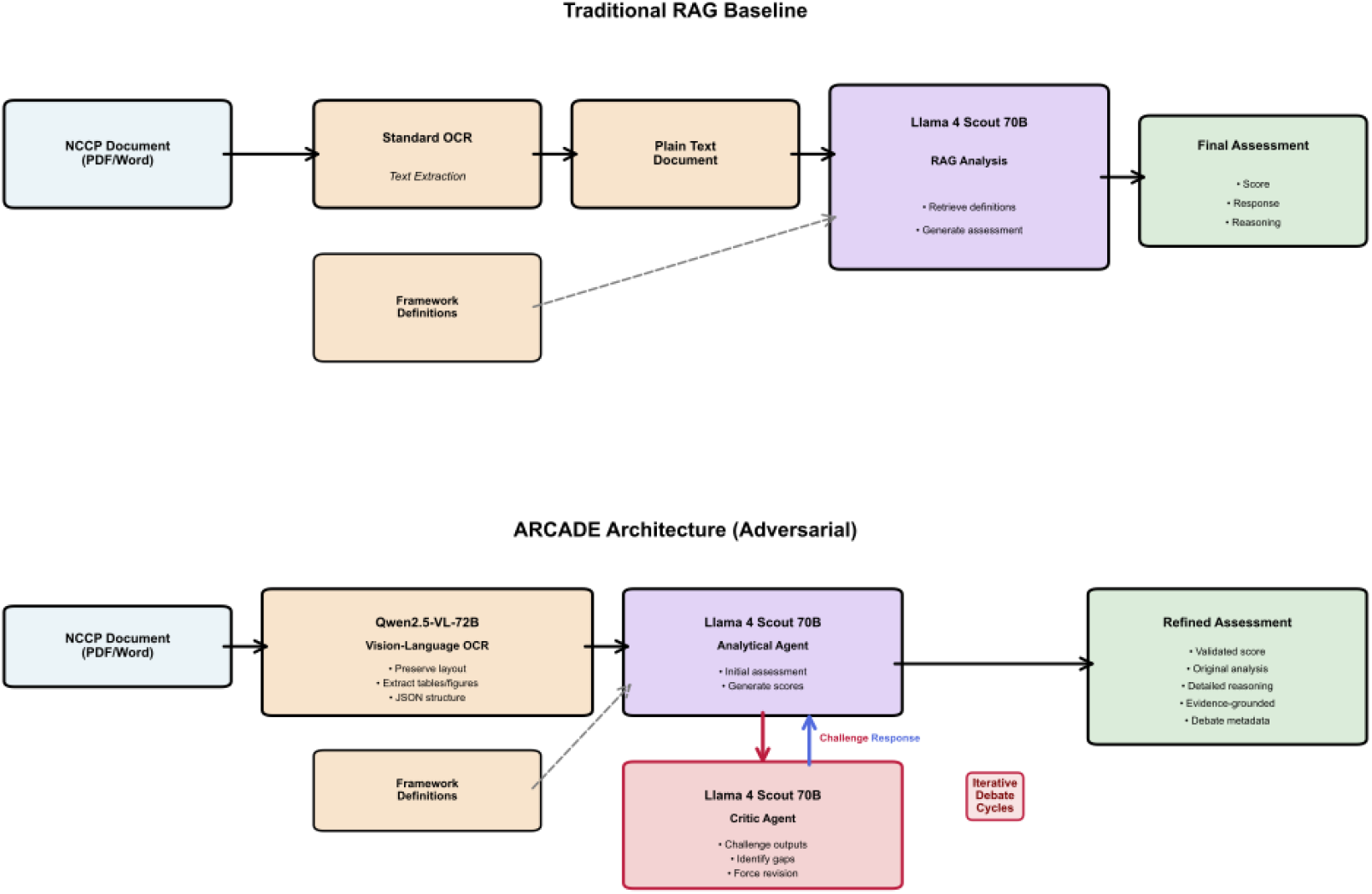
Comparison of Traditional RAG Baseline and Adversarial Multi-Agent Validation Pipelines. The traditional RAG baseline (top) processes documents through standard OCR and single-pass analysis, producing scores without mechanisms for self-critique. The ARCADE architecture (bottom) introduces a dedicated critic agent that challenges initial assessments through iterative debate cycles, producing validated scores with evidence-grounded reasoning. Both pipelines retrieve framework definitions from a shared vector database, but ARCADE’s adversarial loop forces deeper engagement with source documents.

### 2.1 Traditional RAG Baseline

The baseline architecture represents current best practice for framework-based document assessment, combining RAG with standard document processing. Documents undergo OCR to convert PDF content into plain text. The pipeline uses Llama 4 Scout 70B as the core model, retrieving framework definitions and scoring criteria from a vector database to ground generation in expert standards.^[9]^ For each framework element, the baseline produces three outputs: an analytic response, a quantitative score, and a justification. This reflects standard single-pass RAG: outputs are grounded in definitions but lack mechanisms to contest initial judgments.

### 2.2 ARCADE Architecture (Adversarial)

ARCADE retains the baseline’s analytical stage but introduces a second Llama 4 Scout 70B acting as critic. Prompted for structured opposition, the critic interrogates reasoning chains, seeks counterevidence, and flags inconsistencies.^[17]^ Adversarial prompting employs four strategies: devil’s-advocate tests for alternative readings, evidence-gap analysis for overlooked material, semantic coherence checks for consistency, and comparative-robustness tests to explore whether alternative approaches would change scores. When issues are found, the critic issues cited, actionable challenges. The analytical agent must respond with additional evidence, revised scores, or defense of the original judgment, repeating this cycle until convergence.^[16]^

## 3. Evaluation Framework

### 3.1 Dataset

We evaluate ARCADE using 95 policy documents: 50 NCCPs and 45 NCVDCPs.^[8,9,10]^ These represent ideal test cases for four reasons. First, they are structurally heterogeneous (20-100 pages), containing multi-level hierarchies, cross-referencing, and embedded statistical tables. Second, critical information appears in tables, figures, and multi-column layouts. Third, assessment requires a detailed 76 sub-element framework covering epidemiology, service delivery, clinical guidance, financing, and implementation.^[11,12]^ Fourth, they span ten languages, testing linguistic robustness.

The dual-domain dataset allows us to test whether ARCADE’s improvements generalize across different health policy contexts while maintaining structural similarity in document types and framework requirements.

### 3.2 Comprehensive Cognitive Metrics

To evaluate adversarial validation, we employed 18 cognitive metrics across six capabilities, computed separately for each document type (NCCP and CVD), yielding 36 total metric values for cross-domain comparison. This multidimensional framework captures both surface-level characteristics and deeper semantic properties, enabling systematic comparison between Single-Pass RAG and ARCADE.

**Table 1.**
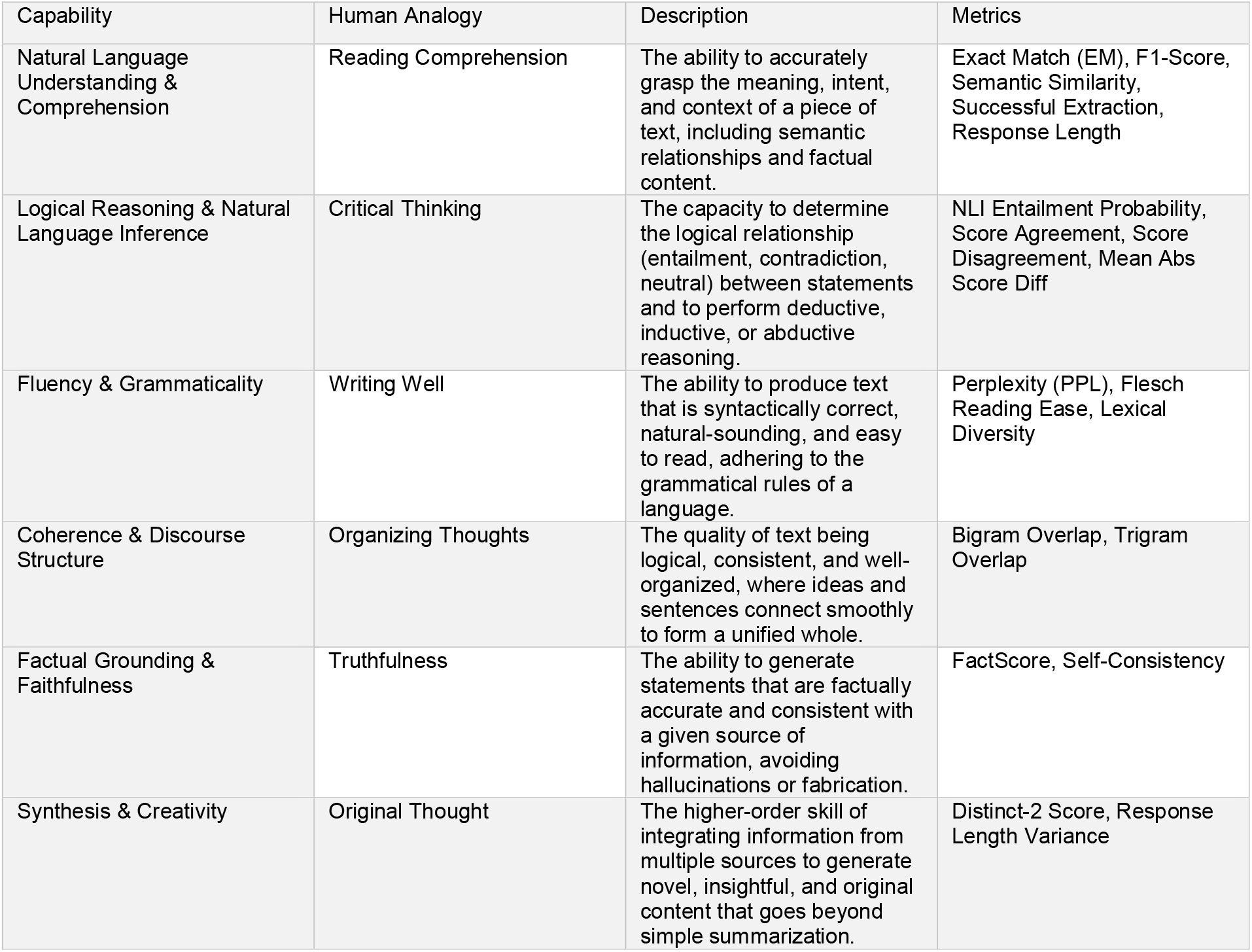
Six-Dimension Evaluation Framework Mapping Human Cognitive Analogies to NLP Metrics. This framework organizes 18 metrics across six cognitive capabilities, each mapped to an intuitive human analogy. Natural Language Understanding assesses information extraction completeness; Logical Reasoning evaluates critical independence; Fluency measures text quality; Coherence captures organizational structure; Factual Grounding verifies accuracy; and Synthesis measures originality. Metrics were computed separately for NCCP and CVD documents, yielding 36 total values.

Natural Language Understanding & Comprehension metrics assess information extraction. Key measures include Successful Extraction Rate (completeness of framework dimension coverage), Response Length (analytical depth), Semantic Similarity (method alignment)^[13]^, F1-Score (token-level overlap), and Exact Match Rate (identical agreement).

Logical Reasoning & Natural Language Inference metrics evaluate critical thinking. Score Disagreement measures the percentage of dimensions where methods assign different scores, indicating independent evaluation. NLI Entailment Probability quantifies logical consistency, while Mean Absolute Score Difference captures the magnitude of scoring divergence.

Fluency & Grammaticality metrics assess text quality and accessibility. Flesch Reading Ease scores readability on a 0-100 scale where higher values indicate greater accessibility.^[14,15]^ Perplexity measures text predictability as a proxy for fluency, and Lexical Diversity (Type-Token Ratio) captures vocabulary variety.

Coherence & Discourse Structure metrics evaluate topical consistency through n-gram overlap. Specifically, Bigram Overlap measures phrase-level consistency, while Trigram Overlap assesses sentence-level coherence.

Factual Grounding & Faithfulness metrics verify accuracy. FactScore quantifies factual overlap by matching entities and numbers between responses^[18]^, while Self-Consistency Score assesses internal factual consistency within responses.^[19]^

Synthesis & Creativity metrics capture originality and variability. Distinct-2 Score measures unique phrase generation, while Response Length Variance quantifies variability in analytical depth across framework dimensions.

## 4. Results

**Figure 2.**
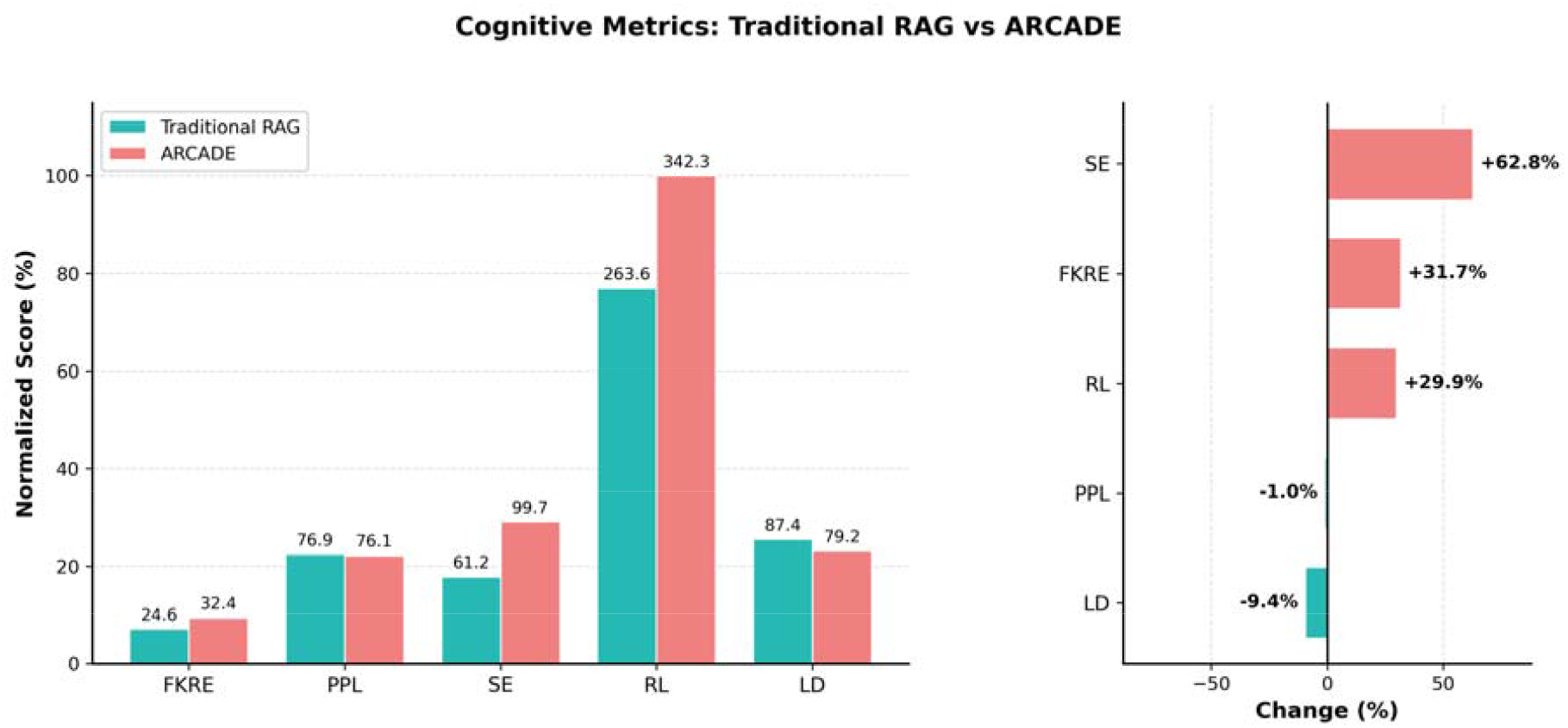
Normalized Performance Scores and Percentage Change Between Traditional RAG and ARCADE. Left panel displays normalized scores (%) for five key metrics: Flesch-Kincaid Reading Ease (FKRE), Perplexity (PPL), Successful Extraction (SE), Response Length (RL), and Lexical Diversity (LD). Right panel shows percentage change, highlighting ARCADE’s substantial improvements in extraction (+62.8%) and readability (+31.7%), with a modest trade-off in lexical diversity (−9.4%). Response length values exceed 100% due to normalization against baseline maximum.

### 4.1 Information Extraction and Comprehension

ARCADE achieves near-complete information extraction across both document types. For NCCPs, successful extraction improves from 59.9% (Single-Pass RAG) to 99.6% (ARCADE), a 39.8 percentage-point gain. NCVDCPs saw similar improvement, rising from 62.9% to 99.7%. This improvement eliminates systematic failures affecting nearly half of the baseline framework dimensions.

Analytical depth also increased substantially: NCCP response lengths grew by 93.4%, and NCVDCP responses by 150.4%. This expansion indicates that adversarial validation prompts significantly more detailed analysis and justification compared to single-pass generation.

**Table 2.** Quantitative Comparison of Single-Pass RAG and ARCADE Across NCCP and CVD Document Types with Effect Sizes. Results are presented for both document types with absolute values, percentage or percentage-point changes, and Cohen’s d effect sizes. Dashes (—) indicate between-method comparison metrics where per-method values are not applicable. Effect size interpretation: |d| < 0.2 = Negligible, 0.2–0.5 = Small, 0.5–0.8 = Medium, ≥ 0.8 = Large. Information extraction and lexical diversity show very large effects in opposite directions, while readability shows a large positive effect.

| Metric | Unit | NCCP<br>Single-Pass | NCCP<br>ARCADE | NCCP<br>Change | CVD<br>Single-Pass | CVD<br>ARCADE | CVD<br>Change | Cohen's *d* |
| --- | --- | --- | --- | --- | --- | --- | --- | --- |
| Information Extraction | % | 59.9 | 99.6 | +39.8pp | 62.9 | 99.7 | +36.9pp | 1.11 |
| Response Length | chars | 200 | 387 | 93.40% | 113 | 285 | 150.40% | 0.16 |
| Flesch Reading Ease | score | 17.9 | 29.9 | 12 | 31.2 | 34.8 | 3.6 | 0.52 |
| Score Disagreement | % | — | — | 60.40% | — | — | 57.70% | — |
| FactScore Overlap | % | — | — | 17.10% | — | — | 30.30% | — |
| F1 Token Overlap | score | — | — | 0.39 | — | — | 0.319 | — |
| Lexical Diversity | % | 83.5 | 75.6 | -9.40% | 91.3 | 82.8 | -9.30% | -1.64 |
| Perplexity | score | 73 | 72.8 | -0.30% | 80.7 | 79.5 | -1.60% | -0.08 |

### 4.2 Critical Independence and Logical Reasoning

Metrics demonstrate genuine critical evaluation rather than redundancy. Score disagreement reached 60.4% for NCCPs and 57.7% for NCVDCPs, indicating independent assessment of over half the framework dimensions. Mean absolute score differences of 1.0-1.2 points (0-5 scale) confirm these disagreements are substantive.

Factual content overlap was limited, with FactScore entity and number matching at 17.1% for NCCPs and 30.3% for NCVDCPs. Similarly, F1 token-level overlap reached only 39.0% for NCCPs and 31.9% for NCVDCPs. These low overlap metrics and high score disagreement establish that ARCADE produces complementary analysis rather than merely rephrasing baseline outputs.

### 4.3 Readability and Accessibility

Despite increased analytical depth, ARCADE substantially improved readability. NCCP Flesch Reading Ease scores rose from 17.9 to 29.9 (+12.0 points), moving from a “very difficult” to “difficult” reading level. For NCVDCPs, scores increased from 31.2 to 34.8. These gains accompanied the 93-150% increase in response length, challenging assumptions that deeper analysis necessarily reduces accessibility.

Perplexity decreased slightly (0.3% for NCCPs, 1.6% for CVD plans), indicating modest improvements in language predictability and fluency. However, lexical diversity decreased by 9.4% for NCCPs and 9.3% for NCVDCPs, suggesting ARCADE employs more consistent but slightly more repetitive terminology.

### 4.4 Topic Coherence and Factual Consistency

Bigram overlap (phrase-level coherence) measured 12.2% for NCCPs and 10.4% for NCVDCPs, while trigram overlap (sentence-level coherence) measured 7.9% and 6.5%, respectively. These moderate levels indicate that Single-Pass RAG and ARCADE maintain some topical consistency while addressing different analytical dimensions.

Self-consistency scores match FactScore values (17.1% for NCCPs, 30.3% for NCVDCPs), confirming internal consistency aligns with between-method factual overlap. Low NLI entailment probabilities (6.9% for NCCPs, 0.1% for NCVDCPs) further demonstrate logical independence between methods.

### 4.5 Synthesis and Response Variability

Distinct-2 scores decreased modestly (0.9% for NCCPs, 1.8% for NCVDCPs), indicating reduced unique phrase generation consistent with the lexical diversity findings. In contrast, response length variance increased dramatically: from 82,023 to 981,897 for NCCPs and from 18,819 to 33,537 for NCVDCPs. This substantial increase suggests ARCADE adapts analytical depth dynamically, providing brief responses for straightforward elements and extensive analysis for complex ones.

### 4.6 Summary of Findings

The comprehensive 36-metric evaluation demonstrates that ARCADE’s adversarial validation architecture produces fundamentally different analytical outputs compared to Single-Pass RAG. Three primary improvements emerge across document types: (1) near-complete information extraction (100% vs. 60%), (2) substantially increased analytical depth (93-150% longer responses) with improved readability (+12 points Flesch Reading Ease for NCCPs), and (3) genuine critical independence evidenced by 60% score disagreement, 17-30% factual overlap, and low token-level similarity. These improvements come with minimal trade-offs: a 9% reduction in lexical diversity suggesting more consistent terminology without sacrificing analytical quality.

**Figure 3.**
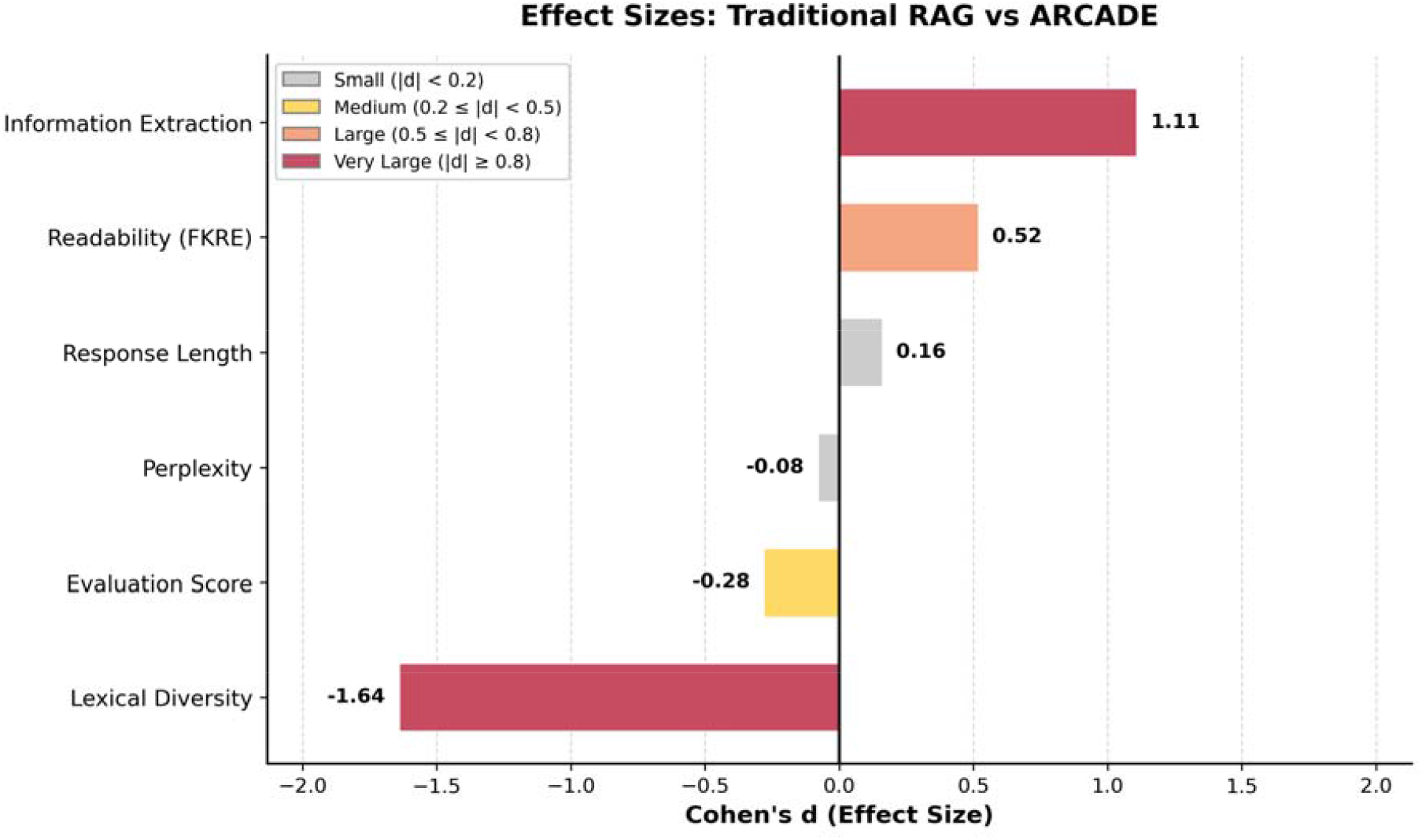
Cohen’s d Effect Sizes Demonstrating Practical Significance of ARCADE Improvements. Horizontal bar chart displaying standardized effect sizes for six key metrics. Positive values (right) indicate ARCADE outperformance; negative values (left) indicate trade-offs. Information extraction (d = 1.11) and readability (d = 0.52) show meaningful improvements, while lexical diversity (d = −1.64) represents the primary trade-off. Perplexity (d = −0.08) and response length (d = 0.16) show negligible to small effects, indicating stability in fluency despite substantially longer outputs.

## 5. Discussion

### 5.1 From Framework Paraphrasing to Original Analysis

ARCADE shifts automated document analysis from paraphrasing frameworks to generating original, document-grounded content. The adversarial mechanism compels the analytical agent to engage deeply with text rather than merely restating framework definitions. Multiple metrics confirm this shift: high score disagreement (60%), low factual overlap (17-30%), and modest token-level similarity (32-39% F1-Score) indicate ARCADE produces complementary analysis addressing distinct aspects of the document.

### 5.2 The Readability Paradox

A counterintuitive finding is the simultaneous increase in analytical depth and readability. ARCADE improved NCCP Flesch Reading Ease scores by 12 points while increasing response length 93-150%. This likely results from the adversarial process, which challenges vague explanations and forces clearer, better-structured arguments. Additionally, the 9% reduction in lexical diversity suggests that using consistent terminology aids readability, albeit at a modest cost to vocabulary variety.

### 5.3 Applications and Future Directions

ARCADE’s success with complex policy documents suggest broader utility in domains requiring framework-based assessment, including regulatory compliance, contract analysis, clinical protocols, strategic planning, and environmental impact analysis. The architecture’s generalization across NCCPs and NCVDCPs indicates applicability to policy documents beyond healthcare.

Future work will explore multi-agent roundtables for structured deliberation and domain-specific fine-tuning to build specialized critic expertise, scaling the adversarial mechanism from bilateral debate to multi-party consultation.

### 5.4 Limitations

Several limitations should be considered when interpreting these findings. First, our evaluation used a single model family (Llama 4 Scout 70B) for both analytical and critic agents. Performance may vary with different foundation models, and the generalizability of ARCADE’s improvements across model architectures remains untested.

Second, our dataset focused exclusively on health policy documents (NCCPs and NCVDCPs). While these represent structurally complex, multilingual documents typical of policy analysis tasks, the architecture’s effectiveness in other domains—such as legal contracts, regulatory filings, or scientific protocols—requires further validation.

Third, the evaluation relied entirely on automated metrics without human expert validation. Although our 36-metric framework captures multiple dimensions of analytical quality, automated metrics may not fully reflect the nuanced judgments that domain experts would provide. Future work should incorporate human evaluation to validate that ARCADE’s outputs meet professional standards for policy analysis.

Fourth, computational costs were not systematically measured. The iterative debate cycles inherent to adversarial validation increase inference time and resource consumption compared to single-pass RAG. For resource-constrained deployments, the trade-off between improved quality and computational overhead warrants consideration.

Fifth, while ARCADE achieved high extraction rates (>99%), we did not evaluate factual accuracy against ground-truth annotations. The metrics capture consistency and independence between methods but do not directly assess whether extracted information correctly reflects document content.

Finally, the assessment frameworks used in this study was developed for cancer and cardiovascular disease control plans specifically. Applying ARCADE to documents with different framework structures or evaluation criteria may require adaptation of the prompting strategies and convergence criteria.

## 6. Conclusion

This work establishes adversarial multi-agent validation as an effective paradigm for automated document assessment. Comprehensive evaluation using 36 cognitive metrics across 6 capabilities demonstrated three primary improvements: near-complete information extraction (100% vs. 60%), substantially increased analytical depth (93-150%) with improved readability (+12 points), and genuine critical independence (60% score disagreement, 17-30% factual overlap). These findings offer significant implications for policy analysis, regulatory compliance, clinical protocols, and any domain requiring rigorous framework-based document assessment.

## Data Availability

All data produced in the present study are available upon reasonable request to the authors.
All data produced in the present work are contained in the manuscript.

